# Characterizing large language model generative artificial intelligence variability in the production of objective structured clinical examination stations

**DOI:** 10.64898/2026.08.04.26359691

**Authors:** Kristen Joseph-Delaffon, Maxime Desgrouas, Sophie Catanese, Julien Lejeune, Julien Nait-Kaci, Eric Piver, Isaure Breteau, Sophie Leducq, Philippe Gatault, Raoul Kanav Khanna, Denis Angoulvant, Nicolas Vallet

**Affiliations:** Neonatology Department, Tours Bretonneau University Hospital, Tours, France; Médecine Intensive Réanimation, Centre Hospitalier Universitaire d’Orléans, Orléans, France; Ophthalmology Department, Tours Bretonneau University Hospital, Tours, France; Pediatric Hematology-Oncology Department, Tours Clocheville Tours University Hospital, Tours, France; Hematology and Cell Therapy Department, Tours University Hospital, Tours, France; Faculty of Medicine & Inserm 1259 MAVIVHe, Tours University, Tours, France; Anaesthesiology and Intensive Care Department, Tours University Hospital,, Inserm 1327, ISCHEMIA, Tours, France; Pediatric Dermatology Unit, Tours University Hospital, Tours, France; Nephrology, Kidney Transplantation and Hemodialysis Department, Tours University Hospital, Tours, France; Ophthalmology Department, Tours Bretonneau University Hospital; INSERM, Imaging Brain & Neuropsychiatry iBraiN U1253, Tours, France; Cambridge Centre for Brain Repair and MRC Mitochondrial Biology Unit, Department of Clinical Neurosciences, Cambridge University, Cambridge, United Kingdom; Faculty of medicine & UMR Inserm 1327 ISCHEMIA, University of Tours, F37000, Tours, France; Hematology and Cell Therapy Department, Tours University Hospital, Inserm 1069, N2COx, France

**Keywords:** Large language models, Objective Structured Clinical Examination (OSCE), Medical education, Generative artificial intelligence, Content validity, Competency-based assessment, AI-generated content

## Abstract

**Background:** Designing high-quality Objective Structured Clinical Examination (OSCE) stations is a time-consuming process. Generative artificial intelligence (AI) represents a promising path to accelerate content creation by automating the generation of scenarios. A growing number of AI tools is now available for this purpose.

**Objective:** To assess the variability between generative AI models in their ability to produce OSCE stations in the field of paediatrics.

**Methods:** A structured prompt was developed based on the French national OSCE guidelines for medical education. Five distinct AI models were provided with this prompt, alongside the neonatal jaundice chapter from the French pediatric reference textbook, to generate 6 complete OSCE stations. *Results.* Prompt compliance was high for ChatGPT 5.1, ChatGPT 5.2, Gemini 3.0 Pro, and Claude Opus 4.5, while it was lower for Grok 4.1. Expert-rated quality was generally high, with few factual errors or missing information across models. However usability differed significantly between models. This was also true for several quality dimensions such as checklist clarity, embedding of checklist answers within vignettes, and ease of standardized patient formation. ChatGPT 5.1 required the most revisions and Gemini most often rated usable as is. Significant inter-model differences were observed in diagnostics, only with ChatGPT 5.1 sampling all three neonatal jaundice categories. Contextual variables showed systematic narrowing across models. Clinical grid density was consistent (10-12 items per station), but thematic distribution differed markedly. Soft skills coverage varied significantly across models (p=0.002), none of them consistently representing all communication competency domains.

**Conclusion:** Large language models can generate structurally compliant OSCE stations, but surface compliance conceals substantive inter-model differences in diagnostic coverage, contextual diversity, and soft skills representation, that compromise content validity. No model currently meets the criteria for unsupervised deployment in a summative assessment bank. The choice of model carries pedagogical implications and expert curation remains essential before integration into high-stakes assessment workflows.

## Introduction

Originally developed in the 1970s, objective structured clinical examinations (OSCE) require students to perform procedural tasks or to interact with simulated patients and peers, allowing for a standardized evaluation of clinical reasoning, communication, and practical skills [1,2]. OSCE have been widely adopted for several decades in the United Kingdom, United States, and Canada as a structured method for training and evaluating the clinical competencies of medical students [3]. Meanwhile, in France, OSCE have recently been included as an assessment component within a national ranking examination governing the allocation of medical specialty and training location.

Despite the pedagogical value of OSCE, they require time and expertise to be built, through designing clinically valid scenarios, elaborating marking grids with minimal inter-subject variability, and piloting stations with standardized patients [4]. The validity of assessment instruments in medical education is commonly evaluated through a framework including content validity, defined as the degree to which an assessment adequately samples the intended construct domain[5]. For OSCE stations libraries, content validity requires that scenarios represent the full range of clinical presentations, patient demographics, and competency domains specified by the curricular blueprint.

Generative artificial intelligence (AI) in the form of large language models (LLMs) such as OpenAI’s ChatGPT [6], Google Gemini [7], Anthropic Claude [8], or xAI Grok [9] are now capable of generating complex, contextualized text with minimal human input. In medical education specifically, these tools have the potential to write OSCE scenarios and evaluation grids. Preliminary evidence suggests that AI-generated content can be leveraged both in the production of OSCE materials and in the formative assessment of student performance [10,11]. Yet, the emergence of AI-generated content may introduce bias, within OSCE station banks, toward certain diagnostic categories, patient profiles, or competency evaluation. In addition, the assessment may fail to meet validity standards regardless of its compliance with formatting requirements.

One source of bias may originate from LLM output variability [12], the evaluation of which should not be viewed as a technical issue. It is rather a validity concern, with direct implications for future OSCE station banks.

To evaluate how LLM variability impacts OSCE station repositories, a comparison was performed qualifying and quantifying the variability of outputs produced by the 5 best-performing LLMs identified during the study period, when tasked with a standardized prompt to write OSCE stations and evaluation grids. The results inform academic physicians on the conditions under which generative AI tools can reliably support assessment in medical education.

## Methods

### Study design

This study used a comparative, cross-sectional design to evaluate outputs of 5 generative AI models tasked with producing OSCE stations in pediatrics. Six stations were generated by each model, yielding a total of 30 stations. All stations were generated on December 15^th^, 2025 **(Figure 1A)**.

**Figure 1.**
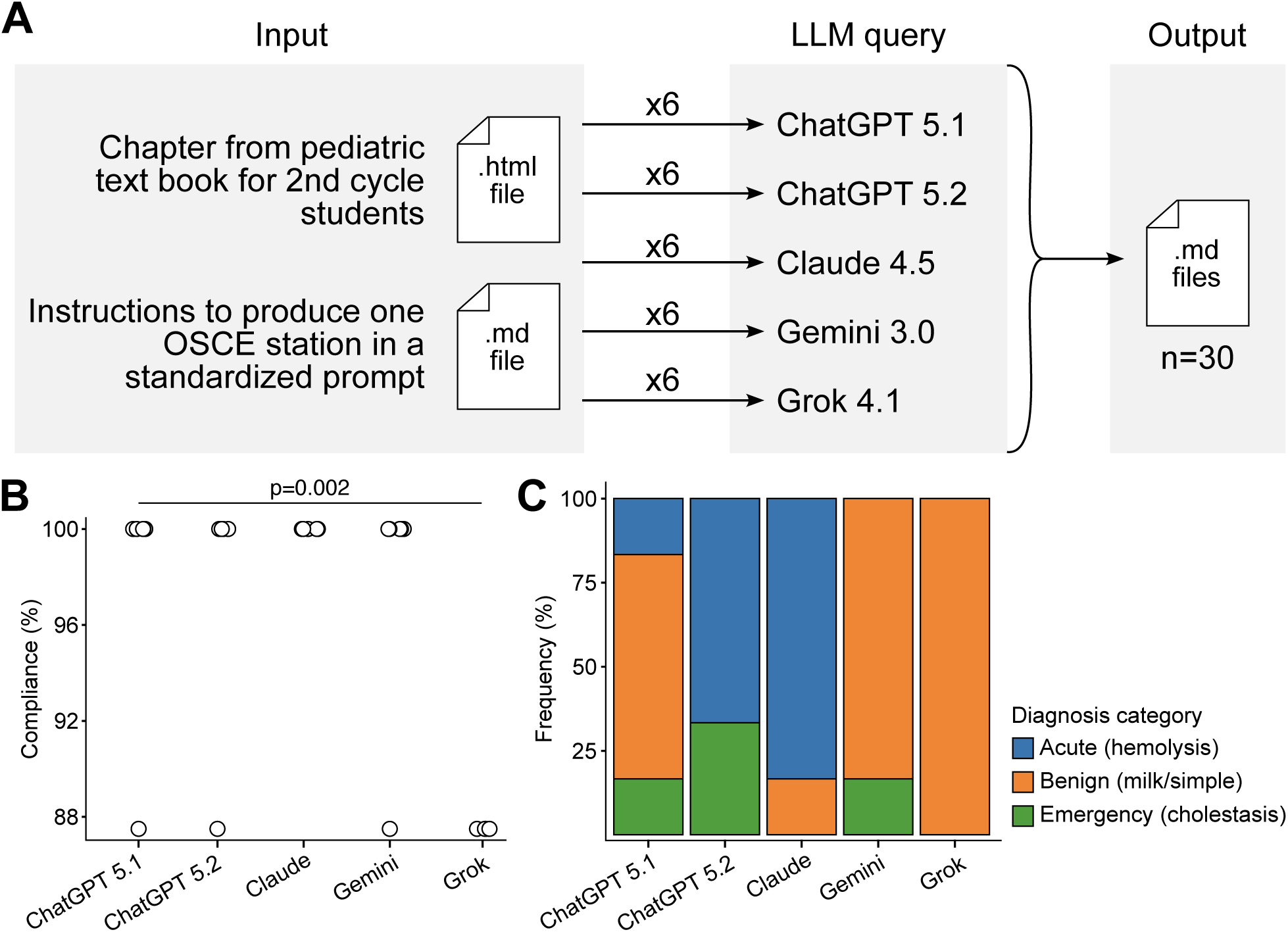
Study overview. A. Method design. The topic is neonatal jaundice and the prompt describes prerequisites asked to the large language model (LLM). Each query was performed on a new LLM session preventing LLM overlap from previous demands. B. Dotplots showing the distribution of compliance to prompt requirements when producing objective structured clinical examination stations (OSCE). Statistical analyses were performed with non-parametric Kruskal-Wallis test. C. Stacked barplots depicting the frequency of diagnosis category among LLM.

### AI models and generation conditions

The 5 AI models selected for comparison were those ranked highest on the openlm.ai Chatbot Arena leaderboard [13] at the time of the experiment. They were respecrively ChatGPT 5.1 Thinking (OpenAI, referred to as ChatGPT 5.1), ChatGPT 5.2 Extended Thinking (OpenAI, referred to as ChatGPT 5.2), Claude Opus 4.5 (Anthropic, with extended thinking enabled, referred to as Claude), Gemini 3.0 Pro (Google, with thinking enabled, referred to as Gemini), and Grok 4.1 (xAI, referred to as Grok) **(Figure 1A)**.

Except for Grok, models were accessed through their paid subscription tiers in order to accommodate the full length of both the reference chapter and the prompt within a single context window.

To ensure the absence of cross-station carryover of outputs, each station was generated within a new conversation session, with memory and data retention explicitly disabled.

For each generation, the model received simultaneously a standardized prompt in Markdown format and the reference chapter in HTML format, and was instructed to produce the station output in Markdown format. These files are available on a dedicated repository: https://doi.org/10.57745/ZCKDJ7.

### Topic selection and reference material

The clinical topic selected was neonatal jaundice, drawn from the official French medical reference book for second-cycle students, publicly available online [14]. Neonatal jaundice was chosen for its narrow and well-codified diagnostic scope. The latter encompasses a small set of benign (physiological, breast-milk) and pathological causes (hemolysis, infection, biliary atresia). This low variability makes any deviation in AI-generated OSCE stations readily detectable. To ensure reproducible input, the corresponding chapter was obtained in HTML format from pediatrician reference book and also available on the dedicated repository: https://doi.org/10.57745/ZCKDJ7 [15].

### Prompt development

A prompt was produced to incorporate all requirements specified in the French national guidelines for OSCE stations in the second cycle of medical education [16]. This prompt, formatted in Markdown, included explicit instructions regarding the structure, content, and format of the expected output.

### Station structure

Each generated station was required to include the following 6 components: (1) a student vignette presenting the clinical scenario, (2) a standardized patient information sheet, (3) an examiner synopsis, (4) a clinical aptitude evaluation grid, (5) a communication and attitudes evaluation grid, and (6) a list of materials required to set up the station.

### Prompt compliance

Compliance with the generation prompt was assessed using 9 binary criteria, evaluating whether each station i)used the required Markdown format, ii)provided a title that did not reveal the diagnosis, iii)included a “you must” and “you must not” instruction section, iv)included a standardized patient scenario, v)included an examiner synopsis, vi)included a clinical aptitude grid, vii) included a communication and attitudes grid, viii)included a global performance section and ix)included a station setup section. An overall compliance score was calculated for each station as the percentage of criteria met.

### Content Variability

The following variables were extracted to assess variability across models: the role assigned to the student, diagnosis to be established, clinical setting, demographic variables (gestational age, neonatal age, birth weight), medical situation variables (breastfeeding status, maternal blood group, parity, infant sex) and standardized patient characteristics (age, sex). Diagnostic outputs were thematically categorized by the principal investigator into three groups: urgent condition (neonatal cholestasis), benign condition (milk jaundice or physiological jaundice), hemolytic condition (group incompatibility).

### Evaluation Grid Analysis

For the clinical aptitude grid, the number of items per station was recorded, and items were thematically categorized by the principal investigator. The number of competency domains assessed, and the types of aptitudes included in the communication and attitudes grid were also analyzed.

### Text Analysis

The clinical context, standardized patient identity, opening sentence, examiner synopsis, and the “you must” and “you must not” sections were quantitatively analyzed. Lexical analysis was performed with word frequency analyzed globally across all models and separately within each model. Comparative word clouds were generated for visual inspection of lexical variation across models.

### Quality assessment

A quality assessment grid was developed based on previous work from Zouakia et al. [11]. The questionnaire included binary questions and Likert scales from 0 to 5 to assess the presence of errors, missing information, clarity, pedagogical validity, educational value, realism, feasibility and usability. All stations generated by AI were evaluated by 4 independent experts used to OSCE, including 2 pediatricians and 2 other practitioners, blinded to the model used.

### Statistical analyses

All analyses were performed using R (version 4.4.0). Given the sample size and the potential non-gaussian nature of data distribution, between-model comparisons of continuous variables were conducted using non-parametric Kruskal-Wallis test. Binary variables were managed using Fisher test. Continuous variables were reported as mean and standard deviation (SD) or median and interquartile range (IQR), and categorical variables as counts and percentages. Content diversity was quantified by counting the number of unique values per model for key variables. A p-value below 0.05 was considered statistically significant. Figures were produced in R with the ggplot2 package and laid out using Inkscape software.

### Ethics considerations

This study did not involve human participants, patient data, nor any identifiable personal information.

## Results

### Prompt Compliance

As indicated, 30 OSCE stations were generated (6 per model). Compliance scores differed significantly across models (*p*=0.002; **Figure 1B**, **Table 1**). ChatGPT 5.1, ChatGPT 5.2, Gemini and Claude achieved median compliance scores of 100%, with minimal intra-model dispersion, indicating adherence to prompt specifications. Grok displayed the lowest median compliance (87.5%). This difference was attributable to a single systematic violation where, contrary to instructions, the station title consistently included the diagnosis.

**Table 1.**
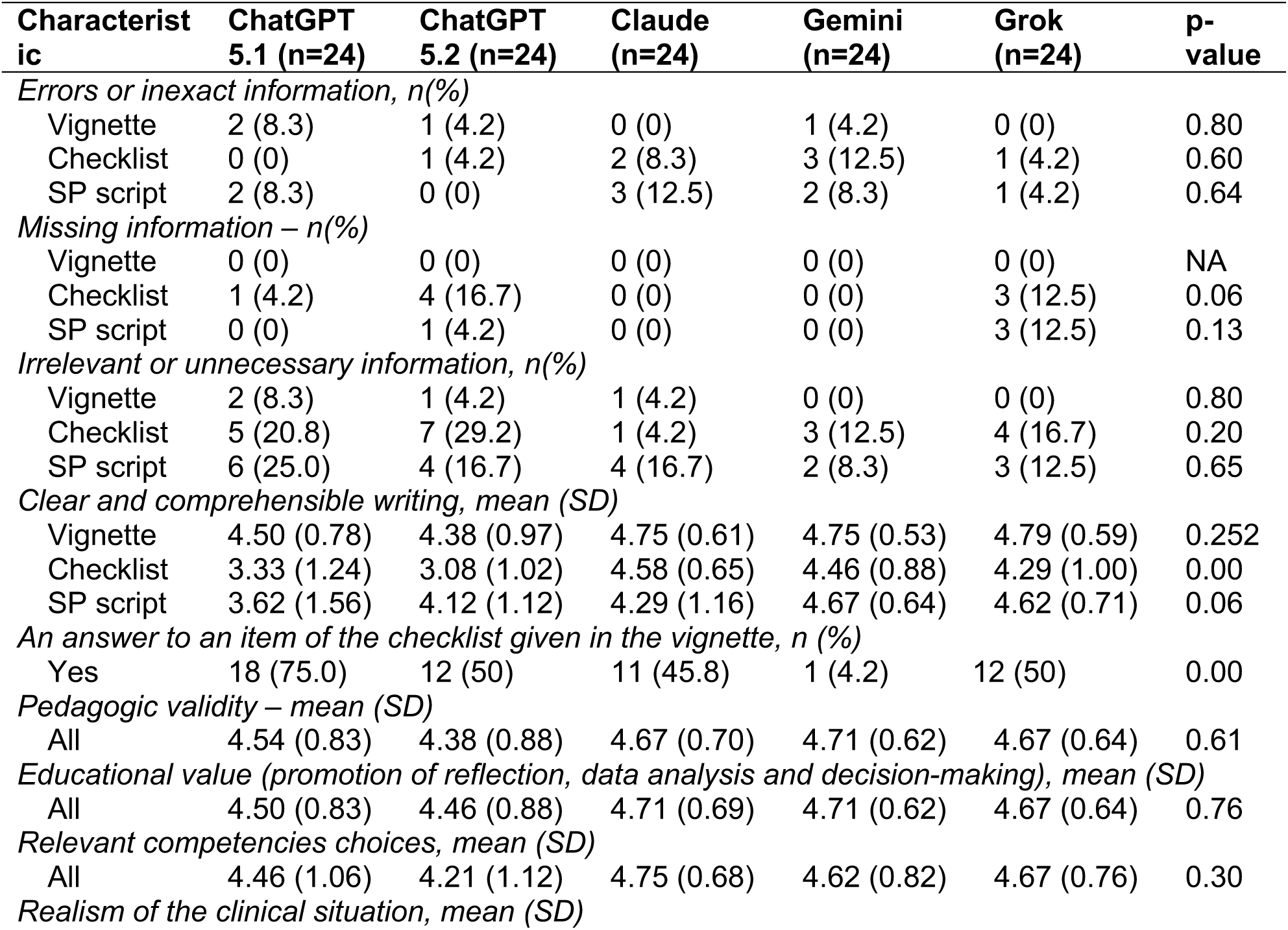
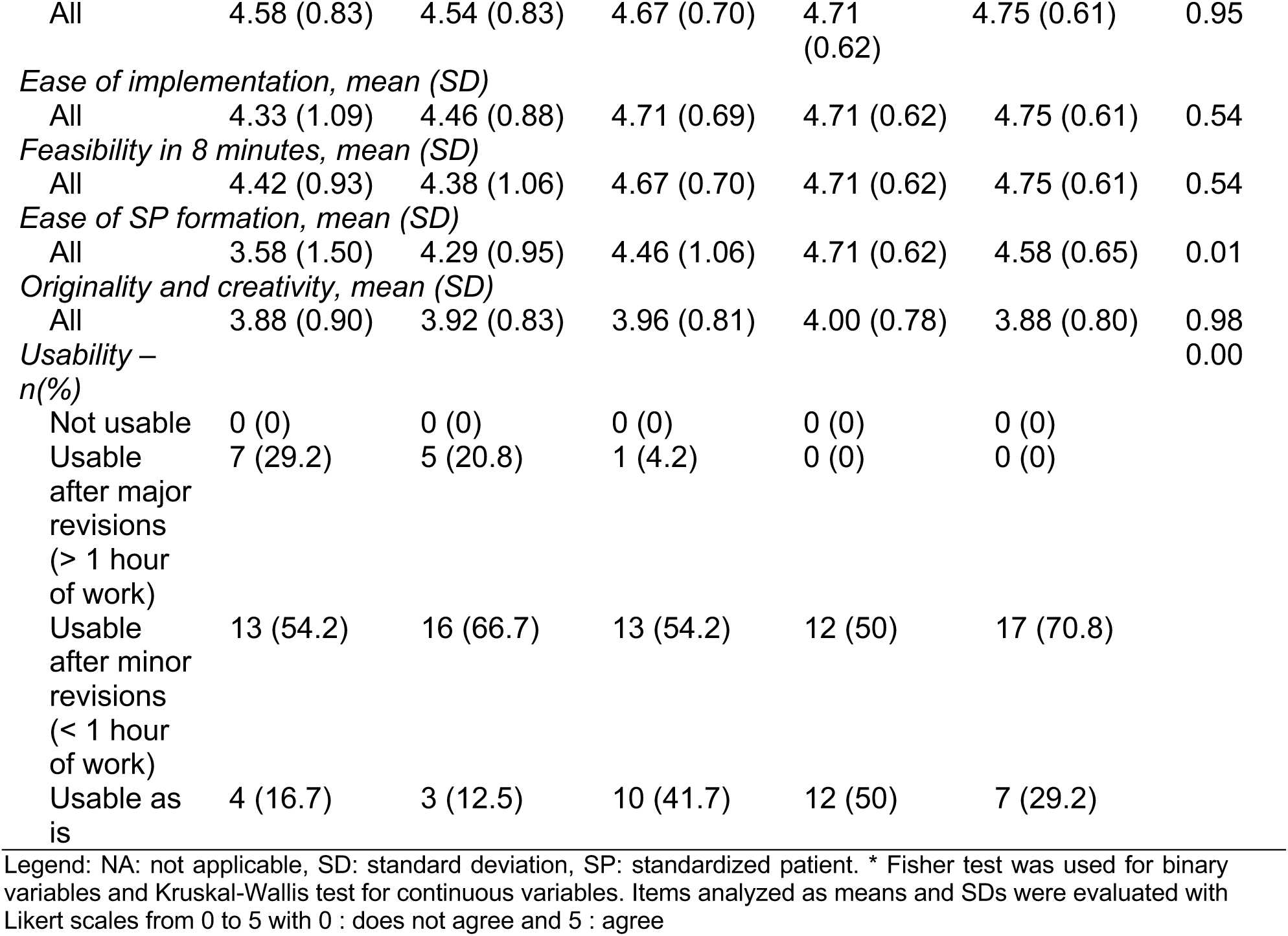
Quality assessment of stations produced by generative artificial intelligence.

### Content variability

#### Diagnostic diversity

Only ChatGPT 5.1 sampled all 3 diagnostic categories defined for neonatal jaundice: benign (n=4/6, 66.7%), acute hemolytic (n=1/6, 16.7%), and cholestatic (n=1/6, 16.7%). Grok and Gemini generated mostly benign presentations (n=6/6, 100% and n=5/6, 83.3% respectively). ChatGPT 5.2 and Claude predominantly produced hemolytic jaundice scenarios (n=4/6, 66.7% and n=5/6, 83.3% respectively, **(Figure 1C)**.

#### Clinical context

All stations featured breastfed infants, and all models assigned the role of the mother to the standardized patient (SP) **(Figure 2A-B)**. Median age of the SP was 29 (IQR: 28-32) years and was comparable between models **(Figure 2C)**. Primiparous scenarios predominated across models, with little diversity in parity.

**Figure 2.**
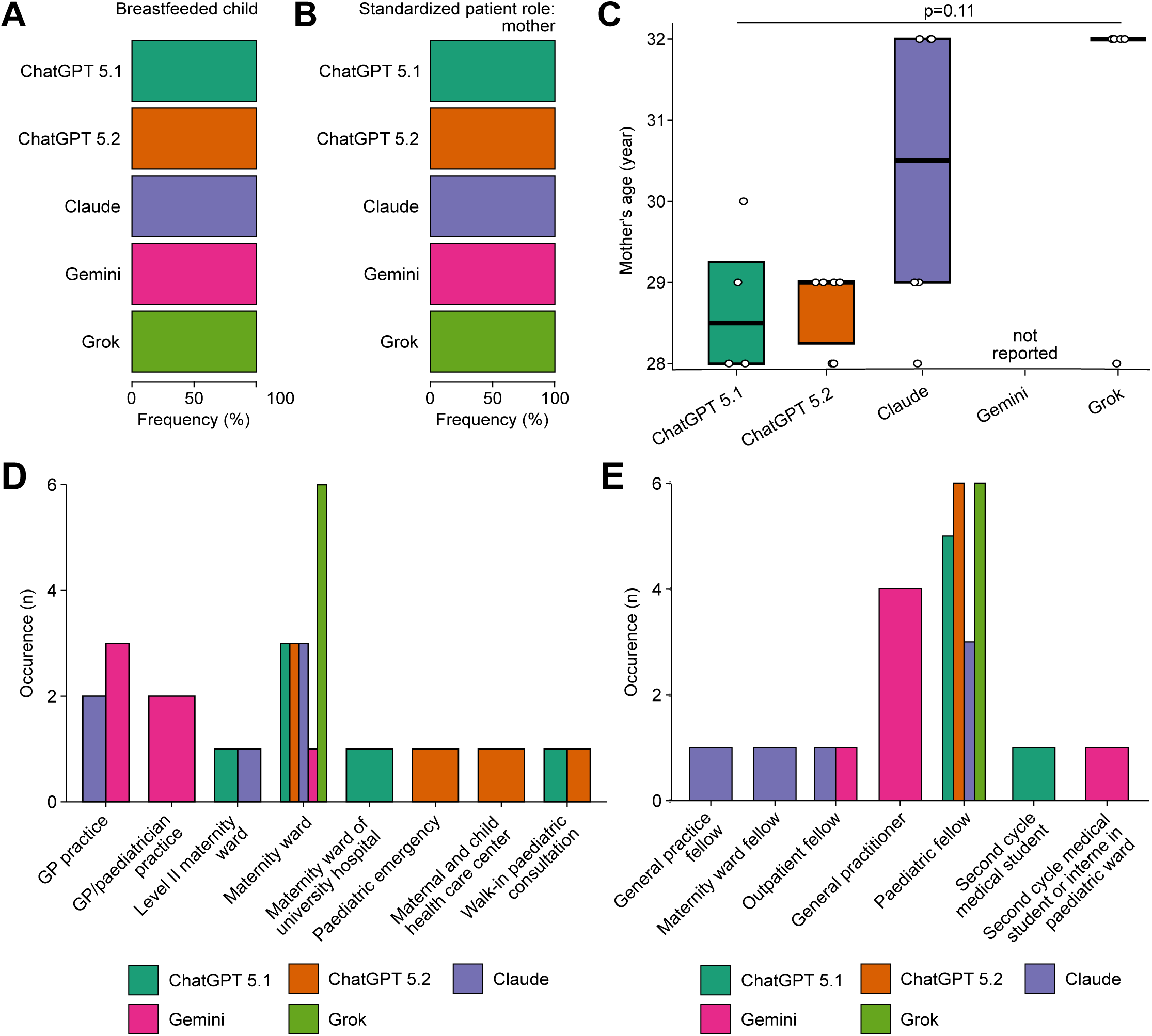
Produced context of the objective structured clinical examination stations (OSCE) A. Barplots showing the frequency of OSCE with a breastfed child B. Barplots describing the frequency of OSCE with mother as a standardized patient role C. Boxplots depicting the distribution of mother age when standardized patient. Statistical analyses were performed with non-parametric Kruskal-Wallis test. D. Barplots illustrating the occurrence of settings of the generated OSCE. GP: general practice E. Barplots showing the occurrence of student roles in the generated OSCE

The settings selected by models spanned a broad range. ChatGPT 5.1 showed a preference for maternity-based scenarios (n=5/6, 83.3%), whereas ChatGPT 5.2, Claude and Gemini produced greater setting diversity (general practitioner office, pediatric emergency ward, pediatric consultation, **Figure 2D**).

The student role assigned similarly varied across models **(Figure 2E)**. Claude generated the broadest range of student roles (pediatric or general practice resident, resident in maternity ward placement or outpatient placement), making its stations more readily adaptable across training levels. ChatGPT models predominantly assigned a single recurring role (pediatric resident, n=5/6, 83.3% for ChatGPT 5.1 and n=6/6, 100% for ChatGPT 5.2).

#### Patient characteristics

Median age of the infant was 3 days (IQR: 3-12), with no significant difference across models (*p*=0.087) except for Gemini which showed a median of 14 days (IQR: 3-18) **(Figure 3A)**.

**Figure 3.**
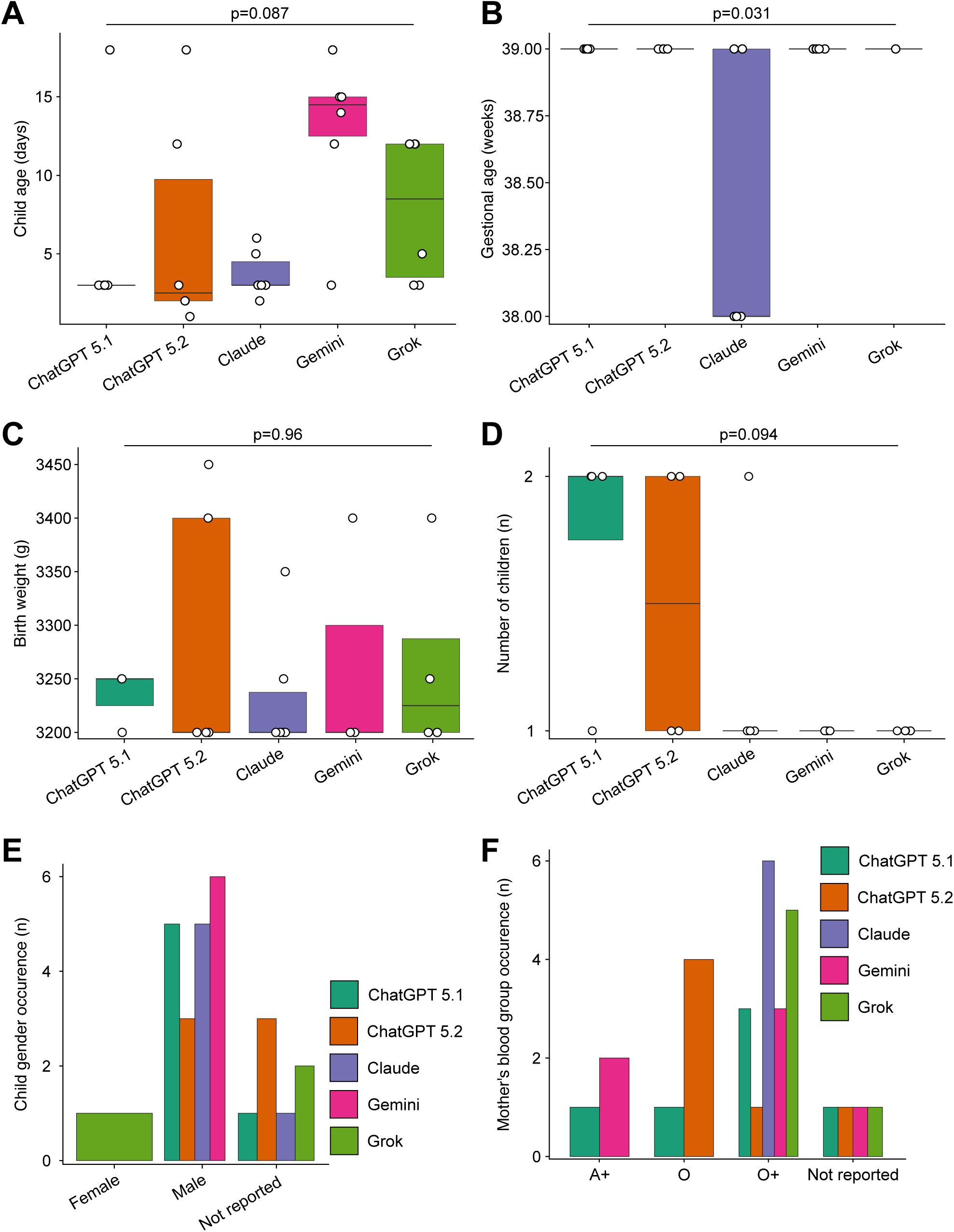
Specific output in the context of the examined chapter. A. Boxplots depicting child age. B. Boxplots showing gestational age. C. Boxplots illustrating birth weight. D. Boxplots displaying the number of children of the mother. E. Barplots illustrating the occurrence of child sex. F. Barplots showing the occurrence of mother blood group. All statistical analyses were performed with non-parametric Kruskal-Wallis test.

Gestational age at birth was significantly lower for Claude (38 weeks, IQR: 38-39) than for ChatGPT, Gemini and Grok (39 weeks, IQR: 39-39; p=0.031; **Figure 3B**). Birth weight, and parity were not significantly different (*p*=0.958, p=0.094, respectively, **Figure 3C-D**).

Child sex **(Figure 3E)** and maternal blood group **(Figure 3F)** were handled differently across models. Claude consistently specified maternal blood group, while it was not always the case for the others (n=5/6, 83.3% for each model). When specified, groups O and O+ were the most commonly assigned maternal blood types (n = 3/6, 50% for Gemini, n=4/6, 66.7% for ChatGPT 5.1, n= 5/6, 83.3% for ChatGPT 5.2 and Grok 4.1 and n=6/6 for Claude), a clinically relevant choice for hemolytic jaundice scenarios, but one that was not always made explicit. For sex, only 1 station from Grok displayed a baby girl (n=1/6, 16.7%). All models prioritized boys (n=3/6, 50% for ChatGPT 5.2 and Grok, n=5/6, 83.3% for ChatGPT 5.1 and Claude, n=6/6, 100% for Gemini) when defined.

The number of unique maternal and infant names per model ranged from 1 to 5 (Figures 12-13). ChatGPT 5.1, Gemini, and Claude generated the greatest diversity (for maternal names, n=4, n=4 and n=5 respectively and for infant names, n=3, n=3 and n=4 respectively). By contrast, ChatGPT 5.2 and Grok produced recurring names across stations (for maternal names, n=2 and n=1 respectively and for infant names, n=3 and n=2 respectively).

#### Stations

The number of items in the “you must” section did not vary much (median of 3 items per station). A similar pattern was observed for the “you must not” section (median=2, **Figure 4A**).

**Figure 4.**
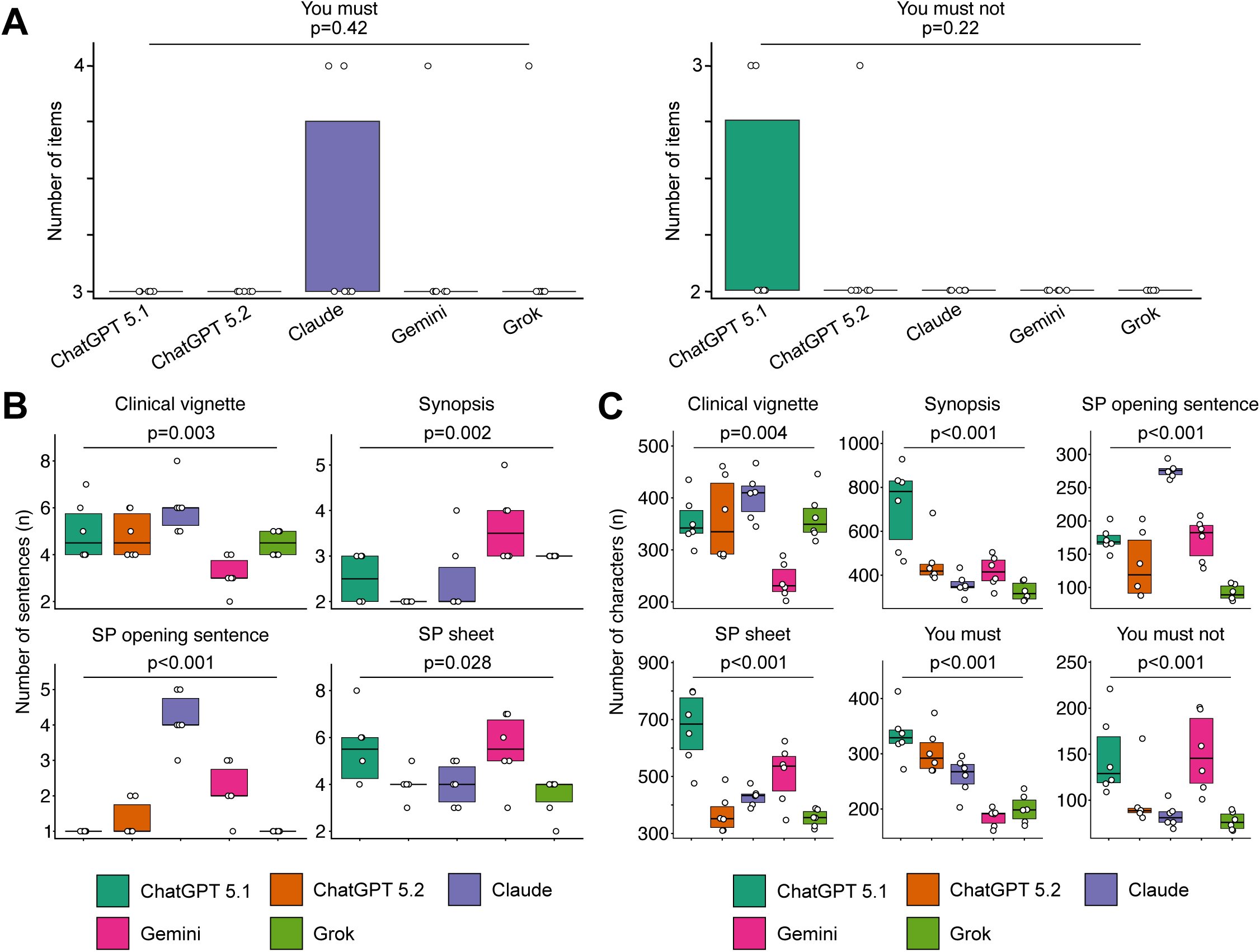
Length of objective structured clinical examination stations sections. A. Boxplots showing the number of items written in “You must” and “You must not” sections B. Boxplot depicting the number of sentences in each section of the vignette C. Boxplot illustrating the number of characters in each section of the vignette All statistical analyses were performed with non-parametric Kruskal-Wallis test.

Considering length and sentence structure, significant inter-model differences in both sentence and character count were observed across all analyzed sections. ChatGPT 5.1 generated the longest content overall, particularly in the SP scenario (median of characters=781, IQR: 463-928) and the examiner synopsis (median of characters=684, IQR: 476-799). Grok consistently produced the shortest outputs **(Figure 4B-C)**.

#### Evaluation grid analysis

The total number of items in the clinical evaluation grid did not differ significantly across models (medians between 11.5 and 12, ranging from 10 to 12, *p*=0.276; **Figure 5A**), with item counts ranging from 10 to 12 across all models.

**Figure 5.**
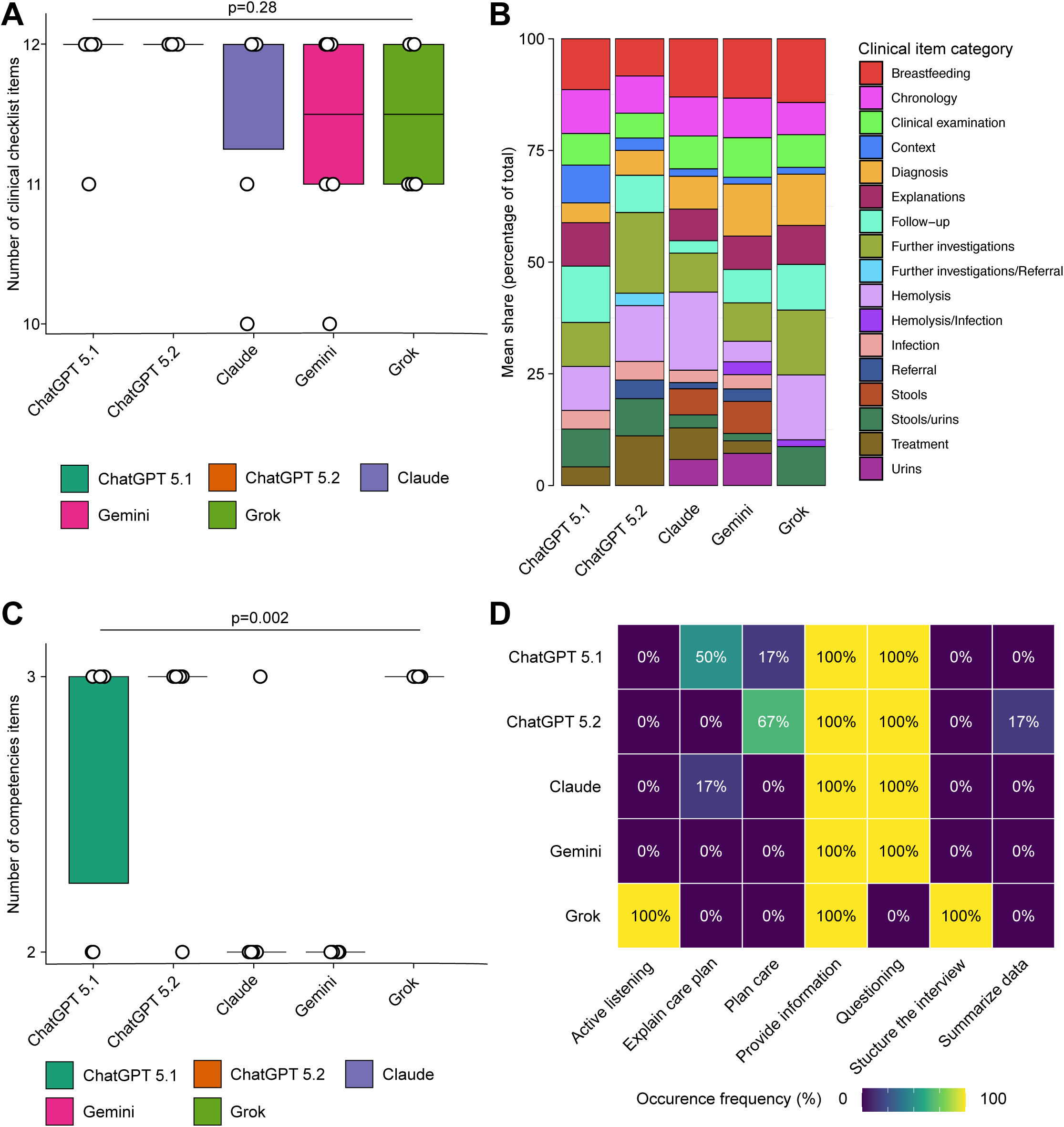
Obtained items in the objective structured clinical examination stations marksheets. A. Boxplots showing distribution of the number of clinical items included in the marksheets. B. Stacked barplots depicting category of clinical items included in the marksheets. The frequency was obtained by calculating the occurrence of each category among the overall number of categories produced by the n=6 replicates. C. Boxplots illustrating the distribution of the number of competence items in the marksheets. D. Heatmap exhibiting the frequency of competences category found in the output marksheets. Frequency refers to the number of occurrences in each replicate among all marksheets from the same large language model. All statistical analyses were performed with non-parametric Kruskal-Wallis test.

The thematic distribution of items within the grid differed markedly **(Figure 5B)**. Seventeen thematic categories were identified. Gemini allocated a larger proportion of items to diagnosis-related categories (11.6%) and breastfeeding (13.3%); ChatGPT 5.2 emphasized diagnostic investigations (18.1%), Claude foregrounded hemolysis-related (17.5%) and breastfeeding items (13%) and ChatGPT 5.1 placed greater weight on follow-up (12.6%) and breastfeeding (11.4%). Grok showed the greatest intra-model variability in thematic distribution (breastfeeding, hemolysis-related items and further investigations between 14.3 and 14.5% each).

Soft skills evaluation was mainly different across models (*p*=0.002; **Figure 5C**), with median counts ranging from 2 (Claude and Gemini) to 3 (both ChatGPT models and Grok). Two competency domains were always represented across models: providing information and questioning/history-taking, except for Grok 4.1 for the latter. By contrast, care planning (ChatGPT 5.1 16.7% and ChatGPT 5.2 66.7%) and management suggestion (ChatGPT 5.1 50% and Claude 16.7%) showed inter-model variability. Active listening and structuring the interview were consistently included in all Grok stations **(Figure 5D)**.

#### Textual analysis

Across all models, the most frequent items in the clinical vignette reflected the shared clinical situation. Beyond this common core, distinct lexical signatures emerged. Claude stood out markedly, with a profile dominated by quantified clinical parameters such as vital signs (min for minutes n=12, fc for cardiac frequency n=6, g for grams n=11, weight n=10), suggesting systematic inclusion of clinical data absent from other models. Gemini produced the most narratively oriented vignettes, with tokens anchored in postnatal follow-up context (“consult” n=5, “days” (of life) n=7). ChatGPT 5.2 emphasized a birth-centered framing (born n=9, term n=5, baby n=6), while ChatGPT 5.1 foregrounded breastfeeding-related items (breastfed n=4, breast n=4) alongside clinical alert markers (fever n=5). Grok displayed the most concise profile, relying on basic demographic (weight n=8, child n=6, mother n=7) and temporal anchors (term n=6, days n=8).

Regarding the SP scenario, character construction strategies diverged substantially. Claude generated the most psychologically nuanced profiles, with tokens evoking maternal guilt (worried n=10). Grok produced the most concise profiles, reduced to structural identification elements (days n=7, role n=7, baby n=8).

Instructional content in the “You must” section shared a common core across models — history-taking, explanation, and further investigations — but diverged in emphasis. Claude prioritized etiological history-taking (anamnestic n=6, information n=6, collect n=6). ChatGPT 5.1 foregrounded diagnostic reasoning (jaundice n=6, pathological n=5). ChatGPT 5.2 broadened the scope towards further targeted investigations (propose n=6, exams n=6). Gemini adopted a diagnostic reasoning framing (orient n=5, interrogation n=6); and Grok oriented directives towards targeted management (propose n=8, adapted n=4).

All models converged on a shared prohibition core in the “You must not” section, centered on performing physical examination and prescribing treatment.

#### Quality assessment

**Table 1** describes the quality assessment filled by the experts. All 5 models produced OSCE stations with few errors (0–12.5% across vignette, checklist, and SP script components), with the highest rates observed for Gemini in the checklist (12.5%) and for Claude in the SP script (12.5%), while missing information was rare overall (≤16.7%). Irrelevant or unnecessary content was frequent in both the checklist and SP script components (up to 29.2%), peaking in the checklist for ChatGPT 5.2. Writing clarity was rated highly for the vignette across all models (mean: 4.38–4.79) but was significantly lower for ChatGPT 5.1 and 5.2 checklists (3.33 and 3.08, respectively) than for Claude, Gemini, and Grok (4.29–4.58; p<0.01). ChatGPT 5.1 most frequently embedded the checklist answer within the vignette itself (75.0%), a pattern significantly less common with Gemini (4.2%; p<0.01).

Pedagogical validity, educational value, relevance of competencies, realism, ease of implementation, and feasibility within 8 minutes were consistently rated higher for Claude, Gemini, and Grok (means: 4.62–4.75) than for ChatGPT 5.1 and 5.2 (4.21–4.58), with the only statistically significant gap seen for ease of standardized-patient formation (ChatGPT 5.1: 3.58 vs 4.29–4.71 for the other models; p=0.01). Originality was comparable across models (3.88–4.00).

Overall usability reflected these trends and differed significantly between models (p<0.01): no station was rated unusable, but ChatGPT 5.1 required major revisions most often (29.2%), whereas Gemini had the highest proportion of stations usable as is (50%) and no station requiring major revisions.

## Discussion

These findings confirm that LLMs can generate formally compliant and clinically plausible OSCE scenarios, but also expose substantial inter-model variability, and random diagnoses selection, raising important concerns about assessment validity and pedagogical equity.

### Comparison with prior work

All 5 LLMs generated broadly usable OSCE stations with low rates of error and missing content, suggesting that generative AI can meaningfully support station drafting for OSCE. However, none of the models was fully error-free or ready-for-use without human oversight. No model had a majority of its stations rated as usable without revision, reinforcing that expert review remains indispensable regardless of the model chosen. Performance nonetheless varied appreciably between models. Claude, Gemini and Grok scored numerically higher than both ChatGPT versions on pedagogical validity, realism and feasibility, although these differences did not reach statistical significance. Conversely, overall usability differed significantly across models, driven by the higher proportion of ChatGPT-generated stations requiring major revisions. ChatGPT 5.1 in particular tended to embed checklist answers directly within the vignette, a flaw that could compromise the discriminant validity of a station by cueing candidates. Both ChatGPT versions also produced significantly fewer clear checklists than the other models, while ease of standardized-patient formation was significantly lower specifically for ChatGPT 5.1. ChatGPT 5.2 performed comparably to Claude, Gemini and Grok on this dimension. Notably, Grok, the only free model, matched or exceeded premium models on several structural dimensions such as realism and feasibility, although it embedded checklist answers within the vignette as often as ChatGPT 5.2, indicating that this specific weakness is not confined to costlier models. These findings should be interpreted cautiously given the single clinical topic evaluated.

Prior studies have shown that GPT-4 was capable of generating OSCE stations with acceptable educational quality when structured prompting strategies are employed with 88% of stations deemed usable without major revision [11]. Here, the results extend this finding to a multi-model comparison, showing that high compliance is achievable across diverse architectures when prompt specifications are sufficiently detailed. Zafar *et al.* identified 10 evidence-informed principles for AI-assisted OSCE design, emphasizing that faculty oversight and structured guidance are prerequisites for safe deployment in high-stakes contexts [17]. Findings of the present study support this position. While all 5 models met formatting requirements, none demonstrated the breadth of diagnostic coverage, contextual diversity, and soft skills consistency that would justify unsupervised deployment in a summative assessment bank. The near-universal prompt compliance observed across models, with the exception of Grok’s systematic inclusion of the diagnosis in station titles, is encouraging from an operational standpoint. It suggests that LLM can reliably follow complex multi-criteria instructions, a prerequisite for any scalable AI-assisted content generation workflow. However, compliance with formatting specifications should not be mixed with educational validity. As previously observed in an experimental study of ChatGPT-4 as an OSCE assessor, surface agreement between AI-generated outputs and expert judgment may be acceptable in some domains (notably history-taking) while remaining poor in others (clinical reasoning, management), with intraclass correlation coefficients far below the thresholds required for high-stakes assessment [18]. Structural compliance also masked substantial inter-model differences in output length across station sections. These differences are not merely stylistic. In the context of OSCE delivery, section length has direct operational consequences. Excessively brief examiner synopses may limit assessor access to the clinical context required for reliable and consistent evaluation of student performance, thereby increasing inter-rater variability. Conversely, excessively detailed standardized patient scenarios threaten to bring cognitive overload for trained actors, potentially reducing the authenticity and reproducibility of the simulated encounter. Optimal section length is therefore a dimension of station quality that should be explicitly specified in generation prompts and verified during expert curation, alongside content and structural criteria.

The most educationally consequent findings of this study concern content variability. The marked disparity in diagnostic coverage across models, ranging from ChatGPT 5.1 coverage of all 3 neonatal jaundice categories to Grok’s exclusive generation of benign presentations. This result highlights potential LLM biases and raises concerns about content validity in AI-generated stations. According to classical test theory, content validity requires that a station bank systematically and representatively samples the full breadth of the targeted construct domain. A bank dominated by a single diagnostic category violates this requirement by construction, irrespective of individual item quality. Beyond this structural failure, uneven diagnostic coverage carries direct implications for the discriminative value of the station bank. In a formative or summative OSCE context, repeated exposure to a single diagnostic category — here, benign neonatal jaundice — introduces predictability effects that may allow students to perform well through pattern recognition rather than genuine clinical reasoning. This undermines one of the core purposes of OSCE design which is to discriminate between students who have achieved the expected level of clinical competence and those who have not. Ensuring diagnostic diversity across a station bank is therefore not merely a matter of content breadth, but a prerequisite for assessment validity.

This concern extends to contextual variables. The repeated assignment of the mother as standardized patient narrows the clinical context of the scenario. This narrowing is reinforced by the predominance of primiparous cases and male infant presentations. Together, these patterns may reduce the validity of the simulated encounter and limit student exposure to the full range of situations they will encounter in clinical practice. While these scenarios individually remain clinically coherent, their systematic uniformity limits the representativeness of the station bank and may reduce its ecological validity across diverse training contexts. A station bank intended for use across multiple institutions, levels of training, or clinical settings requires sufficient contextual variability to ensure that its performance reflects transferable clinical competence rather than familiarity with a narrow and predictable scenario profile. These patterns echo broader concerns about demographic bias in LLM-generated medical content, which have been documented across multiple clinical domains [19,20].

The grid density was broadly consistent across models from 10 to 12 items. This suggests that LLM have followed norms of OSCE scale construction instructed within the prompt. However, the thematic distribution of items differed substantially, reflecting divergent implicit pedagogy. Models that favored diagnosis-related items (Gemini) implicitly signaled a different vision of clinical competence than those emphasizing follow-up (ChatGPT 5.1) or hemolysis-related reasoning (Claude).

LLM failed at including domains such as interview, listening and care planning. Hence, LLM do not apply a consistent model of clinical communication. This is in line with a report of psychometric validation that stated that communication competencies in OSCE stations require careful item construction to achieve acceptable reliability and validity [21]. AI-generated soft skill grids appear insufficiently standardized to meet these requirements without significant expert revision. This inconsistency has a particular impact in the context of competency-based medical education, where OSCE stations are increasingly required to align with validated entrustable professional activities (EPAs). Communication competencies, including active listening, structured information delivery, and shared decision-making, are central to the management of neonatal jaundice. Systematic mapping of AI-generated soft skills grids against validated EPA frameworks should therefore be considered a mandatory step in any quality assurance workflow for AI-assisted station generation. This limitation is compounded by inter-model differences in the psychological and narrative depth of standardized patient profiles. The richness of a simulated patient identity, including family configuration, emotional state, health literacy, and prior experience with the healthcare system, directly shapes the range of communication competencies that can be assessed during the encounter. Models that generated sparse or stereotyped patient profiles therefore constrain not only the realism of the simulated encounter, but also its capacity to assess the full spectrum of relational and communicative skills expected from graduating clinicians. This dimension of station quality remains largely unaddressed in current frameworks for AI-assisted OSCE generation and warrants explicit attention in future prompt engineering and curation guidelines.

The current generation of LLMs occupies an intermediate position in the OSCE design workflow: capable of generating structurally plausible stations, but requiring systematic expert curation before deployment in formative or summative contexts. It can be proposed that this curation should focus on 3 dimensions, respectively diagnostic coverage (ensuring that the station bank samples the full construct domain as defined by the curricular blueprint), contextual diversity (varying setting, student role, patient demographics, and family configuration), and evaluation grid standardization (ensuring alignment of soft skills coverage with validated communication frameworks). This is in line with previous findings which highlighted the importance of teacher intervention for OSCE station quality compared to LLMs [22]. Importantly, the choice of model is not neutral. Results of this study suggest that different models apply different pedagogical orientations, with implications for what students are expected to demonstrate and how examiners are guided to assess them. Lexical analysis provides a window into these divergent orientations that metrics alone cannot capture. The vocabulary deployed across station sections encodes implicit assumptions about what clinical reasoning looks like, which information is salient, and how clinical uncertainty should be communicated. Models that favored quantified, etiologically driven language (emphasizing laboratory thresholds, diagnostic criteria and pathophysiological mechanisms) embodied a vision of clinical competence centered on biomedical knowledge. By contrast, models that produced narratively-rich patient profiles or instructional minimal examiner synopses implicitly prioritized different dimensions of the clinical encounter. These differences are not trivial as the language of a station shapes what students are expected to demonstrate, how examiners are guided to interpret that performance, and ultimately which version of clinical competence is being assessed. Taken together, these lexical findings reinforce the conclusion that AI-generated stations cannot be treated as interchangeable outputs since systematic curation and harmonization of both content and language remains essential before deployment in high-stakes assessment contexts.

### Limitations

Several limitations of this study warrant recognition. First, the analysis was restricted to a single clinical topic (neonatal jaundice), which limits the generalizability of findings to other domains. Still, this topic was selected for its concision and well categorized management. The evaluation of station quality was based on structural and textual analysis rather than psychometric data from actual student performance. Validation studies linking AI-generated station characteristics to assessment outcomes are needed to establish the practical significance of the differences observed. Furthermore, the models compared here represent a specific generation of LLMs assessed at a single point of time. The rapid evolution of models means that findings will require updating as new versions are released. The quality and usability of AI-generated OSCE content from the perspective of faculty evaluators has been addressed elsewhere. Zouakia et al. demonstrated that 88% of GPT-4o-generated stations were deemed usable without major revision when structured prompting strategies were employed [11]. The present study complements this evidence by focusing on inter-model structural and textual variability rather than absolute quality judgement.

## Data Availability

Input files and station outputs are available on the dedicated recherche.data.gouv.fr repository: https://doi.org/10.57745/ZCKDJ7

## Conclusions

This study highlights that LLM are able to produce structurally compliant and clinically plausible OSCE stations. However, the apparent compliance hides substantive intra and inter-model differences. The latter involve diagnostic coverage, contextual diversity, output length, lexical profile and soft skills representation. These compromise content validity, assessment quality and pedagogical coherence. - Currently, full criteria for unsupervised deployment in a summative assessment bank, are not provided by any accessible model. Expert curation remains essential and should focus on 3 dimensions, respectively diagnostic breadth, contextual variability and alignment of evaluation grids with validated communication frameworks and EPA. Moreover, the choice of model is not educationally neutral. Each LLM encodes a distinct pedagogical orientation that determines what students are expected to demonstrate and how examiners assess them. As LLMs become increasingly embedded in medical education workflows, ensuring that their outputs meet the standards of validity, equity, and pedagogical coherence required for high-stakes clinical assessment, must remain a shared priority for educators, assessment specialists, and developers alike.

## Ethics approval statement

This study did not involve human participants, patient data, or identifiable personal information. All Objective Structured Clinical Examination stations were generated by artificial intelligence models from a publicly available clinical reference document and were not used in any real assessment context. No ethical approval was required.

## Acknowledgement

Medical writing for this manuscript was assisted by MPIYP (MC Béné), Paris, France.

## Author contributions

K. Joseph-Delaffon and N. Vallet conceptualized the study.

K. Joseph-Delaffon developed the initial prompt based on M. Desgrouas expertise.

K. Joseph-Delaffon conducted all statistical analyses,

K. Joseph-Delaffon and N. Vallet drafted the first version of the manuscript.

N. Vallet designed the Figures and supervised the study.

K. Joseph-Delaffon, J. Lejeune, J. Nait-Kaci, and S. Catanese evaluated the OSCE stations.

All authors critically reviewed the manuscript and provided their feedback then incorporated by K. Joseph-Delaffon and N. Vallet in the final version.

## Abbreviations

LLM: large-language model
OSCE: objective structured clinical examination
EPA: Entrustable Professional Activities

